# Trustworthiness and Transparency Features Were Less Frequent in Randomized Trials Presenting Large Effects in Abstracts

**DOI:** 10.1101/2025.10.20.25338369

**Authors:** Jonathan F. Henssler, Joana Reis-Pardal, Lina Koppel, John P.A. Ioannidis

**Author notes:** <u>Correspondence to</u>: John P.A. Ioannidis, MD, DSc, 3180 Porter Drive, Stanford Research Park, Room A129, Palo Alto, CA 94304, USA. <u>Conflicts of interest:</u> There are no conflicts of interest.

## Abstract

**OBJECTIVES:** Large effect sizes (ESs), especially when prominently presented in trial abstracts, draw large attention, but it is important to understand whether they are trustworthy. We aimed to assess indicators of transparency and trustworthiness in randomized controlled trials (RCTs) reporting some large ES in their abstract, in comparison with RCTs presenting only non-large ESs in their abstract.

**STUDY DESIGN AND SETTING:** We included RCTs indexed in MEDLINE between January 1, 2024 and March 18, 2025, presenting at least one standardized mean differences of absolute value 0.8 or higher (large ES) versus those presenting only smaller absolute standardized mean differences in their abstract. Trial characteristics and methodological features were extracted systematically in large ES and non-large ES trials. Primary outcome was pre-specified protocol registration, secondary outcomes were having no protocol and public availability or repository placement of raw data.

**RESULTS:** We evaluated 152 trials with large ESs in their abstract and 175 trials with only non-large ESs in their abstract. Large ES trials had suggestively lower rates of pre-registered protocols (45% versus 61%, p=0.0054) and significantly higher rates of no protocol registration (26% versus 13%, p=0.0028) than non-large ES trials. There was no difference in raw data public availability or repository placement (6% versus 7%). Large ES trials were also less likely to be multicenter (p=0.0042), to have high-income country of corresponding author (p=0.0001), to be conducted in high-income country site(s) (p=0.0003), to have a published statistical analysis plan (p=0.0216), and to result from between-group comparisons (p<0.0001). Large effects were significantly more likely to involve non-drug/non-psychological interventions (p=0.0001).

**CONCLUSIONS:** RCTs presenting large ESs in their abstracts are more likely to lack transparency and trustworthiness features and may operate with higher risk of lack of credibility.

**Highlights:**

- This meta-research study assessed the association of effect sizes with features of transparency and trustworthiness of RCTs
- Large effect trials have fewer registered and available protocols
- Studies with large effects sizes are more likely to have a smaller sample size, single center recruitment and provenance from countries without strong clinical research tradition
- Data sharing is poor across all trials regardless of effect size

**What is new?:** *Key findings:*

- This meta-research study indicated that RCTs presenting large effect sizes in their abstracts are more likely to lack features of transparency and trustworthiness.

*What this adds to what is known?:*

- Large effect sizes in abstracts of trials draw attention and may seem as compelling evidence for a study’s claim. Yet there have been divergent findings concerning the association between effect size and credibility of a trial. Our study showed that large effect sizes are consistently associated with a range of methodological shortcomings.

*What is the implication and what should change now?:*

- RCTs presenting large effect sizes may operate with higher risk of lack of credibility.
- Rather than indicating a convincing finding, extreme results should be met with increased attention and careful scrutiny.

## Introduction

Large effect sizes (ESs) in clinical trials draw attention and may be desirable for investigators and journals to publish. However, an important question is whether they can be trusted. A large ES may be too large to be true, i.e. it may be spurious, inflated and occasionally even fraudulent or fake [1–3]. Previous work suggests that in some fields large ES claims are rarely replicated [4]. In an evaluation of over 85,000 Cochrane meta-analyses, effect sizes exceeding 5 in the odds ratio scale were not uncommon, but they were largely observed in very small trials and they largely diminished or completely evaporated when tested in additional larger RCTs [5]. It is also established that abstracts tend to report extreme results [6], as investigators may try to highlight the most prominent findings, further adding favorable spin to the emerging narrative [7,8]. Non-significant results may be presented as significant in the abstract [9] and it is possible that larger ESs may also be selected for showcasing in the abstract. If so, focusing on abstracts might give an enriched sample in claims of large ESs. Abstracts are the most read part of any publication, including RCTs, and they are most influential in shaping perceptions about the results and importance of the presented work. Favorable spin can further shape clinicians’ interpretation of the evidence and thus decision-making [10].

Protocol pre-registration and demonstrable availability of raw data may offer strong support for the trustworthiness and transparency of an RCT [11,12]. Here we aimed to assess empirically whether protocol registration (in particular prospective registration) and data availability are largely absent in trials that prominently report large ESs in their abstracts. For contrast, we assessed how these trials differ versus RCTs that also report numerically on some ES(s) in their abstract, but in which none of the reported ESs is large.

Given that transparency practices of pre-registration and data availability have only recently become modestly common [13], we focused on very recent RCTs. Assessment of the more recent practice may then also offer evidence that is relevant for the current state of RCTs and their potential trustworthiness, whenever they report large ES. We further aimed to explore whether such RCTs have other features that also differentiate them from trials with non-large reported ESs, e.g. small sample size, corresponding author or performance in non-high-high-income countries, single-center design, specific research areas or medical fields, and publication in journals of lower impact.

## Methods

### Protocol

This is a meta-epidemiological assessment with a case-control design of RCTs that present ESs in the form of standardized mean differences (SMDs) their abstract. The protocol of this study has been pre-registered and published at the Open Science Framework (https://osf.io/8xasw). Methods followed the PRISMA reporting guidelines, including the adaptations for meta-epidemiological studies and checklist [14]. The methods are described in further detail in the published protocol. Clarifications to the pre-specified methodology were required during the conduct of the analysis in some cases, as described below.

### Eligible studies

We searched the PubMed database, and selected RCTs published between January 1, 2024 and March 18, 2025, that reported at least one large ES in their abstracts; and a control group of RCTs reporting only non-large ESs in their abstracts. We only included RCTs reporting standardized mean difference (SMD) or related measures, to be consistent with the choice of effect metrics. ESs with a magnitude of SMD, Cohen’s d or Hedges’ g of 0.8 or higher in absolute terms (positive or negative) were regarded as large. For each eligible trial abstract, we selected the largest reported ES in absolute magnitude to represent it.

### Data Extraction

After categorizing the trials based on the magnitude of ESs reported in their abstracts, we extracted pre-specified key methodological characteristics. We assessed a pilot sample of data (40 with large ES and 40 with non-large ES) to inform the construction of the data extraction form and sample size calculations. After discussion, a final list of data items for extraction and analysis was defined. To secure inter-investigator reliability, the data extraction process was independently replicated by additional reviewers for an initial pilot subset of 80 trials (JRP, LK). Any discrepancies between reviewers were resolved through discussion among the research team and a refined data extraction protocol was established. The resulting data extraction protocol was then subsequently applied to the complete dataset of included trials by one reviewer (JH).

### Outcomes and Statistical Analysis

Primary outcome measure was the difference in proportions of prospective protocol registration between large ES and non-large ES groups. Prospective protocol registration (“pre-registration”) was defined as registration within 14 days of onset of study recruitment, as suggested previously [3]. With no given study conduct/recruitment dates and/or no given registration date, studies were classified as not-preregistered. Whenever only month of study initiation or recruitment was given, protocol registration was defined as pre-registered only if registration was no later than within that same month.

In the process of collecting information on prospective protocol registration, some issues arose that required clarifications to be made to our original protocol. We initially collected information of start dates from the published manuscript and extracted information on the protocol registration date as defined in the respective protocol registry database. Any information provided in the original article concerning protocol registration was used to search for and identify the corresponding protocol, i.e. protocol registry, registry number, and/or URL. However, given that in several cases a study initiation or recruitment date was not stated in the published manuscript, we also checked for this information in the trial registries, whenever a registration record was available. The presented analyses include all available information from these sources. When there was a discrepancy between the manuscript and registry information on whether the trial was prospectively registered, we preferred the manuscript information. We also performed a sensitivity analysis where we preferred the registry-listed information and another sensitivity analysis where we considered all these trials with contradictory information to be retrospectively registered. Statistical significance level for the primary outcome was set at α = 0.005, as suggested by Benjamin et al.[15] where p-values less than 0.005 are considered statistically significant and values between 0.05 and 0.005 are considered suggestive.

Secondary outcome measure was the difference in proportions of any protocol registration (prospective or retrospective) and of availability of (deidentified) individual participant data (IPD) (categorized as already publicly accessible now or already posted in a repository), between the large ES and non-large ES groups. We extracted information on data availability as stated anywhere in the published trial manuscript. When public availability or repository placement of data was claimed in the publication, this was checked and verified for each individual case. To be categorized as ‘repository placement of data’, the manuscript had to provide information on the data repository and, when data was not accessible (e.g. without further request), the data repository had to contain at least the information that data of the same exact trial was stored. P-values threshold for secondary outcomes was set at 0.05.

For all categorical outcomes, we estimated risk ratios (RRs) and 95% confidence intervals (CIs) and calculated P-values using Fisher’s exact tests. For continuous outcomes, we estimated median and interquartile ranges (IQRs) and calculated P-values using Mann-Whitney U tests.

Additional analyses explored (using a P-value threshold of 0.05 without multiplicity adjustment) whether large ES and non-large ES groups of RCTs differed in other features that may also reflect trustworthiness and transparency: availability of statistical analysis plan (SAP), corresponding author with any retracted article(s) (identified through Retraction Watch [16]; for authors with common names, publications were cross-referenced between Scopus and Retraction Watch databases to ensure accurate attribution to the specific author [17], any data sharing statement in article, statement of data availability upon request, and statement that data will not be shared. We searched for information on SAP within the trial publication and the protocol registry. The SAP had to be a separate document from the paper and/or a separate explicitly titled SAP after an overall protocol. Having simply a section of “statistical analysis” in the paper or protocol would not qualify. We also examined if groups differed in the following characteristics: sample size, ES referring to between-group comparison of the randomized arms (as opposed to within-group comparison in one of the arms or isolated indeterminate ES, a subset or the total study population), statistical significance of the defining largest ES (i.e. declaration of p<0.05, or verbal quote of significance, or 95% CI excluding the null, in the abstract, referring to the largest ES that represents the RCT), corresponding author from high-income country [18], study conducted in high-income country, multi-center trial, journal impact factor, research intervention category (drug/biologic/vaccine, psychological/behavioral/educational, other (e.g. invasive procedures, physical exercise interventions)).

In subgroup analyses, primary and secondary outcome analyses were stratified by country, multicentricity, research intervention, and journal impact factor (above/below median). Differences in between subgroup ORs of primary and secondary outcomes were assessed using Cochrane’s Q test. Multi-variable logistic regression analysis considered all factors with p<0.05 in univariable associations, in addition to the large/non-large ES group membership in assessing protocol pre-registration and no protocol availability as dependent variables. Additional sensitivity analyses of primary and secondary outcomes considered alternative ES thresholds, i.e. SMD of 0.5, 1.0 and 1.5.

All analyses were conducted using R and R Studio, Version 4.5.1, Posit Software, PBC.

## Results

### Eligible RCTs assessed

The MEDLINE database search retrieved a final set of 327 trials that reported a numerical effect size in their abstract – 152 RCT presenting a large ES (0.8 or higher), and 175 RCT presenting only non-large ES (Figure 1 – Flow diagram).

**Figure 1.**
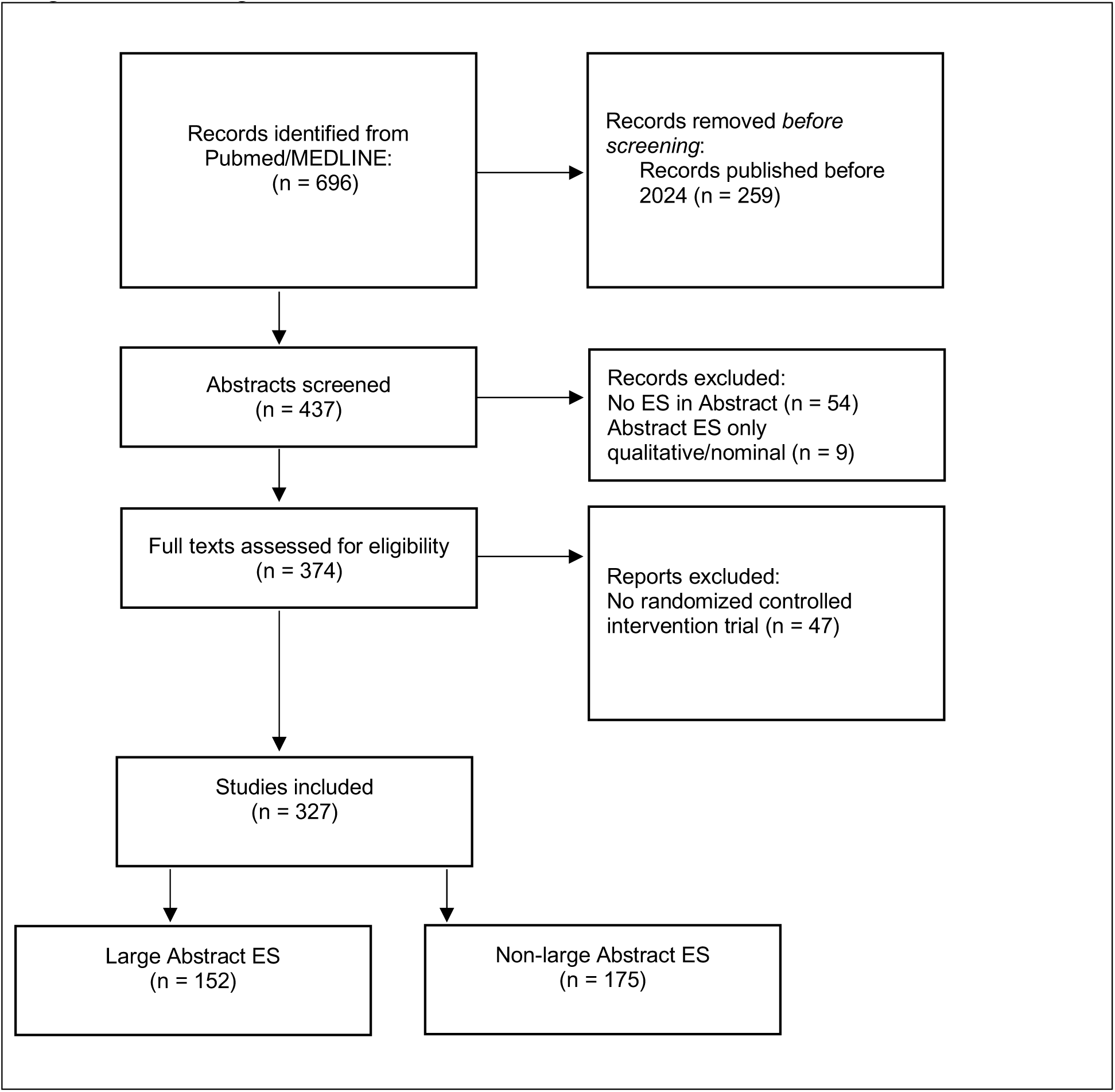
Flow diagram. Identification of records and inclusion of studies – adapted from PRISMA 2020 [19]. ES=Effect size.

All 327 included trials had a median sample size of 92 (IQR: 43.5-191) and a median absolute ES of 0.7 (IQR: 0.5-1.2), when selecting the largest absolute ES reported. Large ES trials had a median sample size of 60 (IQR: 32-110) and a median absolute ES of 1.2 (IQR: 1-1.9). Non-large ES trials had a median sample size of 137 (IQR: 71-261.5) and a median absolute ES of 0.5 (IQR: 0.4-0.6).

### Primary and secondary outcomes

Primary and secondary outcomes are presented in Table 1. There was a suggestion that RCTs reporting some large ES in their abstract were less likely to have a prospectively registered protocol compared with RCTs reporting non-large ES: 45% (95%-CI: 38-53%) versus 61% (95%-CI: 54-68%). The p-value was 0.0054.

**Table 1:**
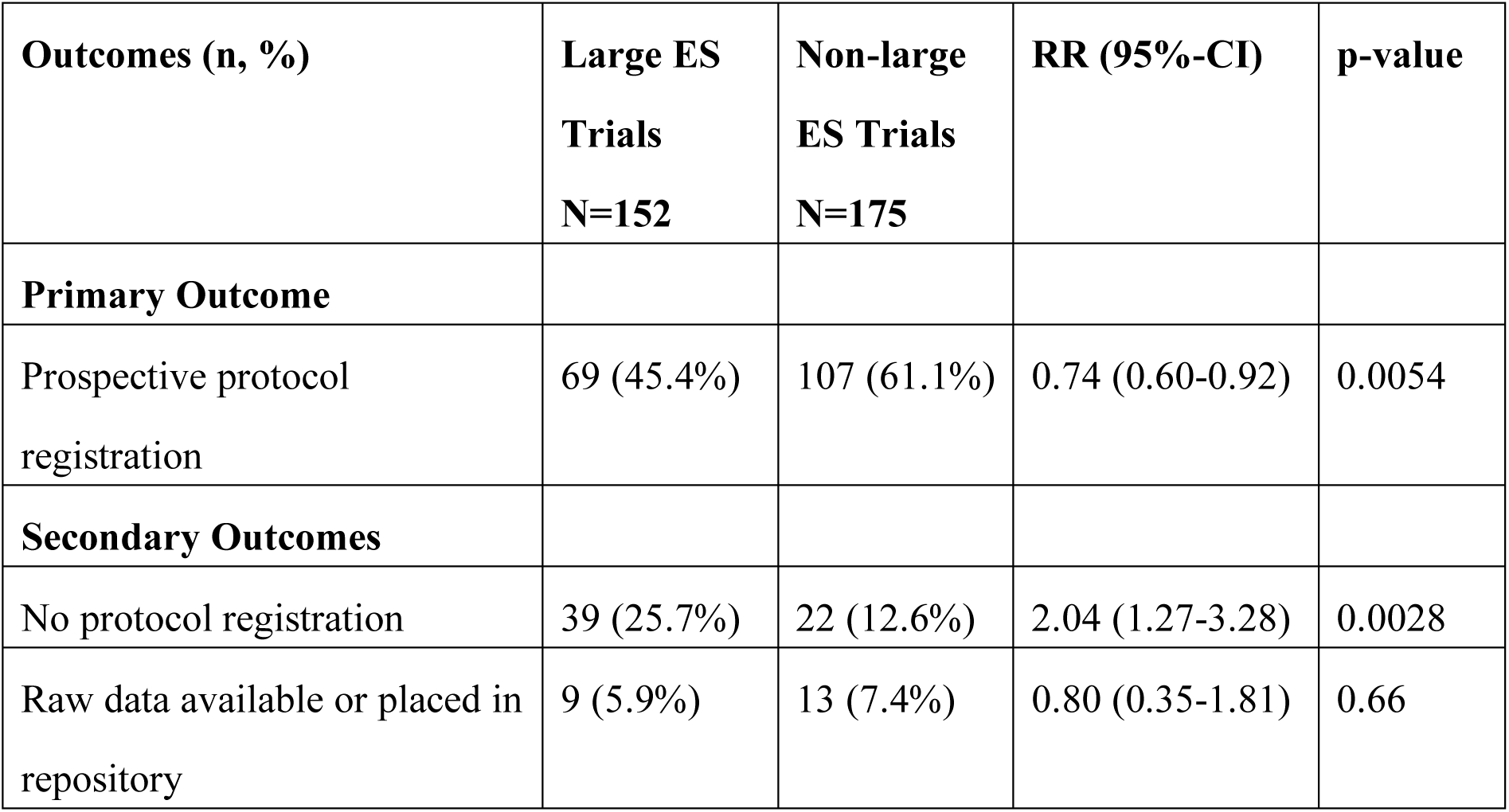
Primary and secondary outcomes. CI=confidence interval, ES=effect size, RR=risk ratio.

| **Outcomes (n, %)** | **Large ES**<br>**Trials**<br>**N=152** | **Non-large**<br>**ES Trials**<br>**N=175** | **RR (95%-CI)** | **p-value** |
| --- | --- | --- | --- | --- |
| **Primary Outcome** |  |  |  |  |
| Prospective protocol registration | 69 (45.4%) | 107 (61.1%) | 0.74 (0.60-0.92) | 0.0054 |
| **Secondary Outcomes** |  |  |  |  |
| No protocol registration | 39 (25.7%) | 22 (12.6%) | 2.04 (1.27-3.28) | 0.0028 |
| Raw data available or placed in repository | 9 (5.9%) | 13 (7.4%) | 0.80 (0.35-1.81) | 0.66 |

RCTs reporting some large ES in their abstract were more likely to have no registered protocol (prospectively or retrospectively) compared with RCTs reporting non-large ES. The proportions of RCTs without any protocol was 26% (95%-CI: 19-33%) in large ES trials versus 13% (95%-CI: 9-18%) in non-large ES trials. The p-value was 0.0028.

Considering either publicly available data or data placed in a repository, overall data availability in included trials was very low and not different (p=0.66) between the two groups. Only 15 trials had readily available raw data (6 in large ES group and 9 in the non-large ES group) and additional 7 trials (3 and 4 in the two groups, respectively) specified that the raw data had been already posted in a declared repository (with the repository containing information on the respective trial).

In sensitivity analyses for the primary outcome, difference in prospectively registered protocol rates amounted to 49% (95%-CI: 41-57%) in large ES trials versus 61% (95%-CI: 53-68%) in non-large ES trials (p=0.034) when preferring registry-listed information, and to 45% (95%-CI: 37-53%) versus 58% (95%-CI: 50-65%) (p=0.020) when considering all trials with contradictory date information to be retrospectively registered.

### Additional outcomes

Additional explorative analyses of methodological features of included trials indicated (Table 2) that RCT reporting large ES in their abstract appeared to differ from non-large ES RCT in several aspects: SAP (p=0.0216), multicenter-trial (p=0.0042), high-income country of corresponding author (p=0.0001), and high-income country conduct site (p=0.0003).

**Table 2:** Additional outcomes. ES=effect size, IQR=interquartile range, JIF=journal impact factor, RR=risk ratio, SAP=statistical analysis plan, s.s.=statistical significance.

| **Features, n (%)** | **Large ES Trials** | **Non-large ES Trials** | **RR (95%-CI)** | **p-value** |
| --- | --- | --- | --- | --- |
| SAP | 6 (3.9%) | 19 (10.9%) | 0.36 (0.15-0.89) | 0.022 |
| Multi-center | 18 (11.8%) | 43 (24.6%) | 0.48 (0.29-0.80) | 0.0042 |
| High-income country author | 94 (61.8%) | 142 (81.1%) | 0.76 (0.66-0.88) | 0.0001 |
| High-income country location | 89 (58.6%) | 135 (77.1%) | 0.76 (0.65-0.89) | 0.0003 |
| Between-group ES | 104 (68.4%) | 156 (89.1%) | 0.77 (0.68-0.86) | <0.0001 |
| Abstract statement on s.s. | 107 (70.4%) | 117 (66.9%) | 1.05 (0.91-1.22) | 0.55 |
| Intervention - Psychological | 78 (51.3%) | 110 (62.9%) | 0.82 (0.67-0.99) | 0.043 |
| Intervention - Drug | 20 (13.2%) | 37 (21.1%) | 0.62 (0.38-1.02) | 0.079 |
| Intervention - Other | 54 (35.5%) | 28 (16%) | 2.22 (1.49-3.32) | 0.0001 |
| Corresponding author has any retracted article(s) - yes | 6 (3.9%) | 6 (3.4%) | 1.15 (0.38-3.49) | 1.0000 |
| **Data sharing features** |  |  |  |  |
| Any data sharing statement | 100 (65.8%) | 125 (71.4%) | 0.92 (0.79-1.07) | 0.28 |
| Access - request to authors | 74 (48.7%) | 90 (51.4%) | 0.95 (0.76-1.18) | 0.66 |
| Explicit statement that data will not be shared | 8 (5.3%) | 9 (5.1%) | 1.02 (0.40-2.59) | 1 |
| Access - data available in public now | 6 (100%) | 9 (90%) | 1.11 (0.90-1.37) | 1 |
| Repository statement - intention | 10 (6.6%) | 20 (11.4%) | 0.58 (0.28-1.19) | 0.18 |
| Repository placement: Repository contains information about the data/study | 7 (77.8%) | 12 (100%) | 0.78 (0.55-1.10) | 0.17 |
| Repository access: Data is accessible on repository now | 5 (71.4%) | 8 (100%) | 0.71 (0.45-1.14) | 0.20 |

Reported effect sizes in large ES trials were significantly less likely to result from between-group comparisons that pertained to the randomized comparison (p<0.0001), and significantly more likely resulted from non-drug/non-psychological interventions (p=0.0001). Furthermore, RCTs reporting large ES in their abstract tended to have lower sample size (median 60 (IQR: 32-110) versus 137 (IQR: 71-262), p<0.0001 by Mann-Whitney U test).

Conversely, journal impact factor did not significantly differ between groups (median 3.8 (IQR: 2.6-4.8) in large versus (4.2 (IQR: 2.8-6)) in non-large ES trials, p=0.101 by Mann-Whitney U test). Also, features of data sharing were similar in both compared groups of trials and data were uncommonly shared (Table 2).

### Subgroup Analyses

Subgroup analyses appear in supplementary Table 3. Results for the primary outcomes were generally not different beyond chance between subgroups, although the difference between large ES and non-large ES trials for no protocol registration was more pronounced in studies with drug or psychological interventions (p=0.022) than in studies with other interventions.

### Regression Analyses

In multivariable logistic regression, only between-group comparison ES was identified as a significant predictor of prospective protocol registration. Studies reporting ES reflecting the comparison between the randomized arms were significantly more likely to have pre-registered protocols (OR = 1.96, 95% CI: 1.11-3.54, p=0.022) (see supplementary Table 4 for all regression analyses results).

### Alternative effect size thresholds (0.5, 1.0, 1.5) for primary outcomes

Prospective registration had suggestive differences between large ES and non-large ES trials even with alternative effect size thresholds: 50% vs 68% (p=0.010), 45% vs 58% (p=0.025), 43% vs 56% (p=0.10) for cut-offs 0.5, 1.0, 1.5, respectively. Respective results for rates of no protocol were 22% vs 9% (p=0.015), 27% vs 15% (p=0.015) and 25% vs 18% (p=0.25). Data sharing was low in all groups regardless of cut-off chosen.

## Discussion

RCTs presenting large ESs in their abstracts had suggestively lower rates of prospective registration and significantly lower rates of any protocol availability compared with studies where all the presented ESs in their abstract were not large in magnitude. Several other features were also associated with reporting large ES, most prominently smaller sample size, lesser likelihood of having multicenter design or SAP, estimating ES from comparisons not protected by randomization, and lesser chance that the corresponding author is from a non-high-income country and/or the trial was done in a non-high-income country. All of these features may be indirect indicators that may herald less trustworthiness and/or transparency and are partially correlated among themselves. Finally, while declarations of intent to share data were common, real public data availability or repository placement were rare for all groups of assessed RCTs.

A previous meta-epidemiological study [20] has also found that across 32 meta-analyses with a total of 165 trials, prospectively registered trials had on average 19% smaller ESs than unregistered ones (relative odds ratio 0.81, 95% CI, 0.65-1.02), while across 37 meta-analyses with a total of 177 trials, those registered at any point had on average 15% smaller ESs than those never registered (relative odds ratio 0.85, 95% CI, 0.67-1.08).

However, these differences were not beyond chance even at traditional p<0.05 thresholds, and most trials in this assessment were fairly old. A smaller meta-epidemiological evaluation of 16 meta-analyses (83 trials) in orthodontics found significantly smaller effects in registered versus unregistered trials (0.36 points in SMD scale) [21]. These finding may reflect that registration may work towards reducing some biases, e.g. those related to selective reporting. Alternatively, registration may be a quality marker that co-exists with other design, conduct and reporting practices that may decrease biases. Specific causal claims should be cautious.

Some other empirical evaluations have noted that non-registered trials may find more often significant, “positive” results and conclusions [22], but this was not an ubiquitous finding, e.g. no such association between registration and positive conclusions has been seen in oncology trials [23] or trials of diverse topics [24].

Previous evaluations used older trials than our evaluation, and registration was less common in the past [25]. Despite increases in the registration rate over time, and also in prospective registration [25,26], many trials are still not prospectively registered. Based on the inconsistencies we noted in manuscripts versus registries, we cannot exclude that additional trials that seem to be prospectively registered may not be so. Lack of proper registration may allow many opportunities for selective reporting and other biases and may facilitate the reporting of large ESs, some of which may not be credible. Previous work has found that registration (and even more so prospective registration) may be associated with lower risk of bias in features like randomization sequence and blinding [27]. Registration well after a study has started or has even been completed remains common [28], but this may not afford similar protection from bias.

Furthermore, even a prospectively registered trial may have many analytical and reporting degrees of freedom, as the registrations may lack sufficient detail. More detail and better rigor may be afforded by the early posting/publication of a full statistical analysis plan [29–34]. Large ES trials seemed to fail more prominently also on this front. Smaller sample size, single center recruitment and provenance from countries without strong clinical research tradition may also increase the chances that bias may be introduced. Disregarding the randomized between-group comparison and presenting within-group effects is a common way to inflate effect sizes. This practice correlated with studies lacking pre-registration or any registration and warrants particular scrutiny.

There is no perfect indicator of trustworthiness for RCTs and all these markers discussed here may be seen as surrogates for protection or susceptibility to diverse biases. Availability of raw data, conversely, may be seen as a more direct measure of transparency. Raw data availability has increased from almost 0% in the beginning of the century to approximately 20% in biomedicine [13]. Progress has been made also for sharing of data from RCTs [35–39], but the rate remains low [38,40]. In our sample of trials, rate of sharing was very low for all trials, regardless of what ESs were reported in the abstract. In the absence of raw data transparency, some authors have raised wider credibility concerns. For example, the experience of John Carlisle as editor of Anesthesia suggested that 30-40% of trials submitted to this respectable journal have been “zombie trials”, i.e. have implausible data that must be either wrong or fake [41]. Progress in data transparency may help alleviate these serious concerns in the future.

Our work has several limitations. First, protocol registration status may only represent an approximation to rigorous methodological process. Protocol detail and requirements of (national) trial registries vary substantially and implementation also varies across trials.

Second, we did not compare methodological detail of protocols to conducted and reported methods in resulting trials. There is large potential for methodological deviations from protocol even with prospective registration [42]. Third, some studies failed to provide recruitment or study initiation dates within the text or provide conflicting dates within manuscript and registry-listed information, carrying a risk of misclassification. Sensitivity analyses generally supported our main findings. Fourth, we focused on trials that use SMD, and our findings may not generalize to other trials, e.g. those with binary metrics. Fifth, reporting of effect sizes in general – and in abstracts in particular – may be more common in specific (sub-) areas of medical research. However, if anything, abstracts that do not report any ESs may be even less rigorous.

Allowing for these caveats, we conclude that RCTs presenting large ESs in their abstracts are more likely to lack features of transparency and trustworthiness. Large ESs in RCTs may superficially seem to be a sign of “success”, but in fact extreme results should be regarded as a sign of possible methodological shortcomings and deserve increased attention and careful scrutiny.

## Contributors

JI had the idea for the study and its design and the other authors contributed to generating the final protocol. JH and JI conducted the literature search and JH screened the articles. JH, JRP and LK reviewed all full texts for inclusion and collected the data. JH analyzed the data. JH and JI drafted the paper and all authors revised the paper and approved the final version.

## Data Availability

The data included in the analyses are available from the authors upon request.

## Acknowledgments

Independent of this work, JH received an outgoing fellowship grant from the Berlin University Alliance and a grant from the German Research Foundation (DFG HE 9191/3-1). <u>Data sharing</u>: The data included in the analyses are available from the authors upon request.

## Supplementary Material

**Supplementary Table 3:** Subgroup comparison analyses for Primary and Secondary Outcomes.

| **Country corresponding author** |  |  |  |  |  |
| --- | --- | --- | --- | --- | --- |
| Prospective Protocol Registration | Subgroup_Variable | Subgroup | OR_95CI | Q_statistic | Q_p_value |
| Subgroup analysis: Protocol Registration by ES split 0.8 stratified by country corr.author | country corr.author == high vs non-high income | high-income group | 0.46<br>(0.27-0.78) | 1.04 | 0.31 |
| Subgroup analysis: Protocol Registration by ES split 0.8 stratified by country corr.author | country corr.author == high vs non-high income | non-high group | 0.78<br>(0.33-1.83) | 1.04 | 0.31 |
| No Protocol Registration | Subgroup_Variable | Subgroup | OR_95CI | Q_statistic | Q_p_value |
| Subgroup analysis: Protocol Registration by ES split 0.8 stratified by country corr.author | country corr.author == high vs non-high income | high-income group | 3.24<br>(1.61-6.52) | 2.99 | 0.08 |
| Subgroup analysis: Protocol Registration by ES split 0.8 stratified by country corr.author | country corr.author == high vs non-high income | non-high group | 1.07<br>(0.38-3.03) | 2.99 | 0.08 |
| Public or repository placed data | Subgroup_Variable | Subgroup | OR_95CI | Q_statistic | Q_p_value |
| Subgroup analysis: public or placed in repository by ES split 0.8 stratified by country corr.author | country corr.author == high vs non-high income | high-income group | 0.74<br>(0.27-2.04) | 0.45 | 0.50 |
| Subgroup analysis: public or placed in repository by ES split 0.8 stratified by country corr.author | country corr.author == high vs non-high income | non-high group | 1.75<br>(0.17-17.49) | 0.45 | 0.50 |
| **Country conduction site** |  |  |  |  |  |
| Prospective Protocol Registration | Subgroup_Variable | Subgroup | OR_95CI | Q_statistic | Q_p_value |
| Subgroup analysis: Protocol Registration by ES split 0.8 stratified by country conduction site | country conduction site == high vs non-high income | high-income group | 0.44<br>(0.25-0.76) | 1.45 | 0.23 |
| Subgroup analysis: Protocol Registration by ES split 0.8 stratified by country conduction site | country conduction site == high vs non-high income | non-high group | 0.79<br>(0.36-1.76) | 1.45 | 0.23 |
| No Protocol Registration | Subgroup_Variable | Subgroup | OR_95CI | Q_statistic | Q_p_value |
| Subgroup analysis: Protocol Registration by | country conduction site == high vs non-high income | high-income group | 3.19<br>(1.55-6.59) | 2.31 | 0.13 |
| ES split 0.8 stratified by country conduction site |  |  |  |  |  |
| Subgroup analysis: Protocol Registration by ES split 0.8 stratified by country conduction site | country conduction site == high vs non-high income | non-high group | 1.25 (0.48-3.29) | 2.31 | 0.13 |
| Public or repository placed data | Subgroup_Variable | Subgroup | OR_95CI | Q_statistic | Q_p_value |
| Subgroup analysis: public or placed in repository by ES split 0.8 stratified by country conduction site | country conduction site == high vs non-high income | high-income group | 0.67 (0.23-2.001) | 0.39 | 0.53 |
| Subgroup analysis: public or placed in repository by ES split 0.8 stratified by country conduction site | country conduction site == high vs non-high income | non-high group | 1.29 (0.23-7.38) | 0.39 | 0.53 |
| **Multicenter Study** |  |  |  |  |  |
| Prospective Protocol Registration | Subgroup_Variable | Subgroup | OR_95CI | Q_statistic | Q_p_value |
| Subgroup analysis: Protocol Registration by ES split 0.8 stratified by Multi- vs. Single-Center formally | Multi- vs. Single-Center formally == multi vs single | multi-center group | 0.22 (0.07-0.70) | 2.88 | 0.09 |
| Subgroup analysis: Protocol Registration by ES split 0.8 stratified by Multi- vs. Single-Center formally | Multi- vs. Single-Center formally == multi vs single | single-center group | 0.65 (0.40-1.06) | 2.88 | 0.09 |
| No Protocol Registration | Subgroup_Variable | Subgroup | OR_95CI | Q_statistic | Q_p_value |
| Subgroup analysis: Protocol Registration by ES split 0.8 stratified by Multi- vs. Single-Center formally | Multi- vs. Single-Center formally == multi vs single | multi-center group | 3.75 (0.87-16.10) | 0.49 | 0.49 |
| Subgroup analysis: Protocol Registration by ES split 0.8 stratified by Multi- vs. Single-Center formally | Multi- vs. Single-Center formally == multi vs single | single-center group | 2.15 (1.15-4.048) | 0.49 | 0.49 |
| Public or repository placed data | Subgroup_Variable | Subgroup | OR_95CI | Q_statistic | Q_p_value |
| Subgroup analysis: public or placed in repository by ES split 0.8 stratified by Multi- vs. Single-Center formally | Multi- vs. Single-Center formally == multi vs single | multi-center group | 0 (0-NaN) | NA | NaN |
| Subgroup analysis: public or placed in repository by ES split 0.8 stratified by | Multi- vs. Single-Center formally == multi vs single | single-center group | 0.79 (0.32-1.98) | NA | NaN |
| Multi- vs. Single-Center formally |  |  |  |  |  |
| **JIF above-below median (3.8)** |  |  |  |  | Q_p_value |
| Prospective Protocol Registration | Subgroup_Variable | Subgroup | OR_95CI | Q_statistic |  |
| Subgroup analysis: Protocol Registration by ES split 0.8 stratified by JIF above-below median (3.8) | JIF above-below median (3.8) == low vs above | low group | 0.81 (0.43-1.53) | 3.01 | 0.08 |
| Subgroup analysis: Protocol Registration by ES split 0.8 stratified by JIF above-below median (3.8) | JIF above-below median (3.8) == low vs above | above group | 0.37 (0.20-0.68) | 3.01 | 0.08 |
| No Protocol Registration | Subgroup_Variable | Subgroup | OR_95CI | Q_statistic | Q_p_value |
| Subgroup analysis: Protocol Registration by ES split 0.8 stratified by JIF above-below median (3.8) | JIF above-below median (3.8) == low vs above | low group | 1.72 (0.81-3.64) | 1.55 | 0.21 |
| Subgroup analysis: Protocol Registration by ES split 0.8 stratified by JIF above-below median (3.8) | JIF above-below median (3.8) == low vs above | above group | 3.68 (1.44-9.42) | 1.55 | 0.21 |
| Public or repository placed data | Subgroup_Variable | Subgroup | OR_95CI | Q_statistic | Q_p_value |
| Subgroup analysis: public or placed in repository by ES split 0.8 stratified by JIF above-below median (3.8) | JIF above-below median (3.8) == low vs above | low group | 2.11 (0.38-11.90) | 1.66 | 0.20 |
| Subgroup analysis: public or placed in repository by ES split 0.8 stratified by JIF above-below median (3.8) | JIF above-below median (3.8) == low vs above | above group | 0.55 (0.18-1.65) | 1.66 | 0.20 |
| **Intervention 'other' (versus psychological or drug)** |  |  |  |  |  |
| Prospective Protocol Registration | Subgroup_Variable | Subgroup | OR_95CI | Q_statistic | Q_p_value |
| Subgroup analysis: Protocol Registration by ES split 0.8 stratified by Intervention | Intervention == other vs drug/psychological | drug/psy group | 0.54 (0.32-0.90) | 0.22 | 0.64 |
| Subgroup analysis: Protocol Registration by ES split 0.8 stratified by Intervention | Intervention == other vs drug/psychological | other group | 0.69 (0.27-1.72) | 0.22 | 0.64 |
| No Protocol Registration | Subgroup_Variable | Subgroup | OR_95CI | Q_statistic | Q_p_value |
| Subgroup analysis:<br>Protocol Registration by<br>ES split 0.8 stratified by<br>Intervention | Intervention ==<br>other vs<br>drug/psychological | drug/psy<br>group | 3.43<br>(1.69-6.98) | 5.27 | **0.02** |
| Subgroup analysis:<br>Protocol Registration by<br>ES split 0.8 stratified by<br>Intervention | Intervention ==<br>other vs<br>drug/psychological | other group | 0.79<br>(0.28-2.22) | 5.27 | **0.02** |
| Public or repository<br>placed data | Subgroup_Variable | Subgroup | OR_95CI | Q_statistic | Q_p_value |
| Subgroup analysis: public<br>or placed in repository by<br>ES split 0.8 stratified by<br>Intervention | Intervention ==<br>other vs<br>drug/psychological | drug/psy<br>group | 0.95<br>(0.36-2.54) | 0.32 | 0.57 |
| Subgroup analysis: public<br>or placed in repository by<br>ES split 0.8 stratified by<br>Intervention | Intervention ==<br>other vs<br>drug/psychological | other group | 0.5<br>(0.07-3.75) | 0.32 | 0.57 |
| **Between-group ES** |  |  |  |  |  |
| Prospective Protocol<br>Registration | Subgroup_Variable | Subgroup | OR_95CI | Q_statistic | Q_p_value |
| Subgroup analysis:<br>Protocol Registration by<br>ES split 0.8 stratified by<br>between-group ES | between-group ES<br>== between vs<br>within/other | between<br>group | 0.58<br>(0.35-0.95) | 0.20 | 0.66 |
| Subgroup analysis:<br>Protocol Registration by<br>ES split 0.8 stratified by<br>between-group ES | between-group ES<br>== between vs<br>within/other | within/other<br>group | 0.75<br>(0.26-2.23) | 0.20 | 0.66 |
| No Protocol Registration | Subgroup_Variable | Subgroup | OR_95CI | Q_statistic | Q_p_value |
| Subgroup analysis:<br>Protocol Registration by<br>ES split 0.8 stratified by<br>between-group ES | between-group ES<br>== between vs<br>within/other | between<br>group | 2.92<br>(1.48-5.77) | 3.53 | 0.06 |
| Subgroup analysis:<br>Protocol Registration by<br>ES split 0.8 stratified by<br>between-group ES | between-group ES<br>== between vs<br>within/other | within/other<br>group | 0.81<br>(0.25-2.56) | 3.53 | 0.06 |
| Public or repository<br>placed data | Subgroup_Variable | Subgroup | OR_95CI | Q_statistic | Q_p_value |
| Subgroup analysis: public<br>or placed in repository by<br>ES split 0.8 stratified by<br>between-group ES | between-group ES<br>== between vs<br>within/other | between<br>group | 0.74<br>(0.27-2.02) | 0.14 | 0.70 |
| Subgroup analysis: public<br>or placed in repository by<br>ES split 0.8 stratified by<br>between-group ES | between-group ES<br>== between vs<br>within/other | within/other<br>group | 1.2<br>(0.12-12.31) | 0.14 | 0.70 |
| **SAP** |  |  |  |  |  |

| Prospective Protocol Registration | Subgroup_Variable | Subgroup | OR_95CI | Q_statistic | Q_p_value |
| --- | --- | --- | --- | --- | --- |
| Subgroup analysis: Protocol Registration by ES split 0.8 stratified by SAP | SAP == yes vs. no | SAP group | 0.59<br>(0.37-0.93) | 1.17 | 0.28 |
| Subgroup analysis: Protocol Registration by ES split 0.8 stratified by SAP | SAP == yes vs. no | no SAP group | 0.19<br>(0.03-1.41) | 1.17 | 0.28 |
| No Protocol Registration | Subgroup_Variable | Subgroup | OR_95CI | Q_statistic | Q_p_value |
| Subgroup analysis: Protocol Registration by ES split 0.8 stratified by SAP | SAP == yes vs. no | SAP group | 2.22<br>(1.24-3.97) | NA | NA |
| Subgroup analysis: Protocol Registration by ES split 0.8 stratified by SAP | SAP == yes vs. no | no SAP group | NA | NA | NA |
| Public or repository placed data | Subgroup_Variable | Subgroup | OR_95CI | Q_statistic | Q_p_value |
| Subgroup analysis: public or placed in repository by ES split 0.8 stratified by SAP | SAP == yes vs. no | SAP group | 0.60<br>(0.23-1.58) | 3.57 | 0.06 |
| Subgroup analysis: public or placed in repository by ES split 0.8 stratified by SAP | SAP == yes vs. no | no SAP group | 9 (0.65-125.32) | 3.57 | 0.06 |

ES=effect size, JIF=journal impact factor, NA=not applicable, OR=odds ratio, SAP=statistical analysis plan.

**Supplementary Table 4:** Multivariable Regression Analysis.

| **Prospective Protocol Registration** |  |  |  |  |  |
| --- | --- | --- | --- | --- | --- |
| Predictor | Coefficient | Std Error | Odds Ratio | 95% CI | P-value |
| Intercept | -0.15 | 0.46 | 0.86 | (0.35 - 2.12) | 0.74 |
| Sample Size (per 500 participants) | -0.041 | 0.05 | 0.96 | (0.81 - 1.04) | 0.41 |
| Single-center | -0.38 | 0.31 | 0.68 | (0.37 - 1.25) | 0.22 |
| Author Country: non-high income | -0.20 | 0.64 | 0.82 | (0.22 - 2.87) | 0.75 |
| Site Country: non-high income | 0.26 | 0.62 | 1.30 | (0.39 - 4.71) | 0.67 |
| Between-group Design (vs Within-group) | 0.67 | 0.30 | 1.96 | (1.11 - 3.54) | **0.02** |
| Intervention: other | -0.40 | 0.37 | 0.67 | (0.32 - 1.38) | 0.27 |
| Intervention: psychological | -0.07 | 0.32 | 0.92 | (0.48 - 1.72) | 0.79 |
| Effect Size Group: non-large ES | 0.43 | 0.24 | 1.54 | (0.95 - 2.49) | 0.08 |
| **No Protocol Registration** |  |  |  |  |  |
| Predictor | Coefficient | Std Error | Odds Ratio | 95% CI | P-value |
| Intercept | -1.16 | 0.61 | 0.32 | (0.09 - 1.002) | 0.06 |
| Sample Size (per 500 participants) | 0.07 | 0.05 | 1.07 | (0.98 - 1.29) | 0.19 |
| Single-center | 0.16 | 0.41 | 1.18 | (0.54 - 2.78) | 0.70 |
| Author Country: non-high income | -0.23 | 0.73 | 0.80 | (0.21 - 3.93) | 0.75 |
| Site Country: non-high income | 0.41 | 0.71 | 1.51 | (0.32 - 5.56) | 0.56 |
| Between-group Design (vs Within-group) | -0.64 | 0.33 | 0.53 | (0.28 - 1.03) | 0.05 |
| Intervention: other | 0.44 | 0.46 | 1.55 | (0.64 - 3.98) | 0.34 |
| Intervention: psychological | 0.0008 | 0.43 | 1.001 | (0.45 - 2.43) | 1.00 |
| Effect Size Group: non-large ES | -0.25 | 0.37 | 0.78 | (0.38 - 1.68) | 0.51 |

ES=effect size.

## Abbreviations

ES: Effect Size
IQR: Interquartile Range
JIF: Journal Impact Factor
OR: Odds Ratio
RCT: Randomized Controlled Trial
RR: Risk Ratio
SAP: Statistical Analysis Plan
SMD: Standardized Mean Difference

